# Burden of renal anemia in 204 countries and territories, 1990-2021 : results from the Global Burden of Disease Study 2021

**DOI:** 10.1101/2024.03.12.24304162

**Authors:** Feifan Chu, Jinzhong Ji, Yuning Ma, Qing Guan, Lumin Chen, Zujie Chen, Qiwei Ji, Mingxin Sun, Hui Zhang, Tingyang Huang, Haihan Song, Xiuquan Lin, Hao Zhou

## Abstract

Renal anemia has inflicted a certain degree of loss on global health. However, there are no systematically analyses on the burden of renal anemia.

We analyzed global prevalence and years lived with disability (YLDs) of renal anemia from 1990 to 2021. Based on the Socio-Demographic Index (SDI), combined with cross-national inequality analysis, frontier analysis and age-period-cohort (APC) model analysis, the prevalence and YLDs differences between different aspects were compared.

Since 1990, the prevalence and YLDs of renal anemia in 2021 have shown an upward trend. In countries with medium and low levels of development, the burden of renal anemia is particularly severe among people over 60 years. A series of analyses based on the SDI revealed a negative correlation between the prevalence of renal anemia and SDI.

Thus, Public health policies targeting renal anemia should give priority to the elderly in medium- and low-development areas.

## Introduction

Renal anemia is a common complication of chronic kidney disease (CKD) and is caused to a certain extent by the reduced production of erythropoietin due to severe renal impairment^[1]^. The occurrence of renal anemia can further worsen the condition of CKD^[2]^ and exacerbate cardiovascular complications associated with CKD^[3,4]^, posing a serious threat to patient safety.

Studies have noted that the global prevalence of CKD stages one to five is 13.4%, and for stages 3-5, the prevalence is 10.6%^[5]^. In a study encompassing 19 CKD cohorts from around the world, 41% of 209,311 patients were reported to have concurrent anemia^[6]^. According to an analysis from a study in the United States, which covered five million insured patients, male severe anemia incidence (hemoglobin < 10 g/dL) increased from 1.3% to 29.7% as the estimated glomerular filtration rate (eGFR) dropped from 60-74 to below 15 mL/min/1.73 m^2^, while in females it increased from 1.9% to 37.6%^[4]^. A multicenter cross-sectional study from China showed a similar trend^[7]^. One of the World Health Organization’s (WHO) global nutrition targets is to reduce the anemia prevalence in women of reproductive age (fifteen to forty-nine years) by 50% by 2030^[8]^. GBD 2021’s latest research has updated and expanded the global estimate of the anemia burden but does not detail renal anemia^[9]^. Although various countries and regions have developed guidelines related to renal anemia^[10,11]^, the elderly, as the main patients with CKD and the main affected subjects of renal anemia, have not received enough attention.

The 2021 Global Burden of Disease, Injuries, and Risk Factors Study (GBD) provides valuable resources for epidemiological research^[9]^. Data obtained from GBD can be used for describing disease burden and analyzing trends, evaluating the current situation and temporal changes of diseases among different populations in various regions and countries, thereby providing information for the formulation of related medical practices and health policies^[9]^.

This study utilized data from the GBD 2021 to describe trends in the age-standardized prevalence rates (ASPR) and age-standardized YLDs rates of renal anemia globally, regionally, and nationally. Additionally, we conducted an international inequality analysis using the standard health equity analysis methodology recommended by the WHO. Furthermore, cutting-edge analysis identified the achievable lowest ASPR and age-standardized YLDs rates for renal anemia in regions of different developmental statuses. Lastly, we applied the APC model to analyze the prevalence and YLDs of renal anemia at the global, regional, and national levels from 1990 to 2021.

## Methods

### 1. Data Source

GBD 2021 offers a comprehensive assessment of health loss due to 371 diseases, injuries, and risk factors across 204 countries and territories, leveraging the latest epidemiological data available. In our study, we extracted estimates of age-standardized prevalence, age-standardized YLDs, and their 95% Uncertainty Intervals (UI) for renal anemia, sourced from the global data, continental divisions (Africa, America, Asia, Europe), and countries (204 in total) from the GBD 2021 website (https://vizhub.healthdata.org/gbd-results/). Our study adheres to the Guidelines for Accurate and Transparent Health Estimates Reporting (GATHER).

### 2. Disease Burden Description

Our research delves into the age-standardized prevalence and YLDs burden of renal anemia across different genders, ages, countries, and regions, as well as the trends between 1990-2021.

### 3. Epidemiologic Transition and Annual Change Rate

Through analysis of the relationship between the burden of renal anemia (age-standardized prevalence and YLDs) in each region and the Socio-demographic Index (SDI), we explored the ties between renal anemia and social development. We projected the expected age-standardized prevalence and YLDs of renal anemia for each SDI value and calculated the observed-to-expected ratios for the burden of renal anemia in each region, identifying countries with significantly higher or lower renal anemia burdens relative to their socioeconomic development levels.

### 4. Rank Change of Renal Anemia

To reflect the shifting burden of renal anemia between 1990-2021, we conducted an in-depth analysis of the rank change of renal anemia among various anemia causes, aiming for an improved representation of the burden of renal anemia.

### 5. Cross-national Inequality Analysis

We employed the Inequality Slope Index and the Concentration Index to estimate renal anemia health inequalities among nations from 1990 to 2021. The Inequality Slope Index was calculated by regressing the age-standardized prevalence or YLDs rates of all age groups across countries against relative positions related to SDI, where relative positions are defined by the midpoint of the cumulative population ranges of SDI rankings. A weighted regression model was used to analyze heteroscedasticity. The Concentration Index was derived by numerical integration of the area under the Lorenz Concentration Curve, fitted to the cumulative scores of age-standardized prevalence or YLDs rates and to the cumulative relative distribution of the population sorted by SDI^[12]^.

### 6. Frontier Analysis

To assess the performance of age-standardized prevalence and YLDs in countries or regions with different levels of development, we conducted a frontier analysis based on age-standardized prevalence, YLDs and SDI using data from 1990 to 2021. The effective gap representing the distance from actual to frontier values, was calculated to highlight the disparity between observed prevalence or YLDs rates and those potentially achievable in a country or region, based their SDI values^[13]^.

### 7. APC Analysis

Using the APC model framework, we analyzed time trends in prevalence and YLDs rates across age, period, and birth cohorts. To prepare input data for the APC model, estimates of renal anemia prevalence, YLDs rates and population data for each country/region from GBD 2021 were used. In the APC model, age effects are represented by fit longitudinal specific-age rates for a given number of birth cohorts adjusted for the period bias. Period/cohort effects are represented by relative rates of prevalence or YLDs during each period/cohort, calculated as the ratio of prevalence or YLDs rates for specific ages in each period/cohort relative to a reference period/cohort. The choice of the reference period/cohort is arbitrary and does not affect interpretation of the results. Two-sided p-values of less than 0.05 were considered to indicate significant differences. All analyses and visualizations were performed in R (V.4.2.1)^[14]^.

## Results

### (1) Age and Gender Trends in Renal Anemia

Regardless of gender, the prevalence and YLDs rates of renal anemia increase with age, with the growth rate accelerating after age 65.

In terms of numbers, the number of cases and YLDs of renal anemia first increase and then decrease, with the highest values appearing between the ages of 70 and 80. Among children under 15, the number of cases and YLDs values are similar between genders, while in other age groups, females have higher numbers and YLDs values than males. This phenomenon is more pronounced in women of childbearing age (15-49 years) and in the elderly over 80 years old. As for prevalence and YLDs rates, the values are relatively close between genders across all age groups (figure 1A, B) (Supplementary Table S1, 2).

**Figure 1.**
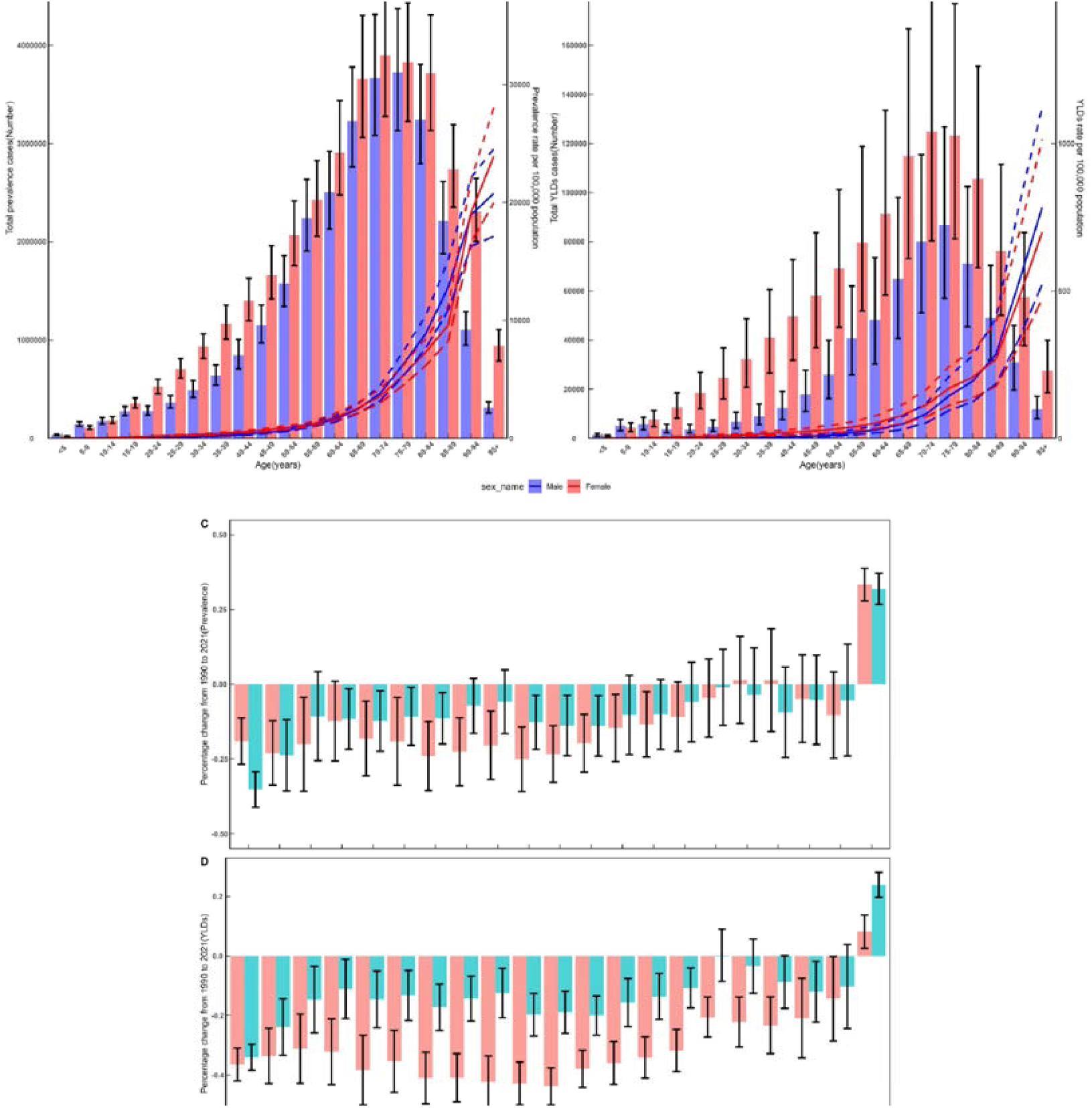
Global prevalence and years lived with disability (YLDs) of renal anemia by age and sex, 2021. (A) Count and Rates of Global Renal Anemia Cases by Age and Sex in 2021, per 100,000 People; (B) Count and Rates of Renal Anemia YLDs by Age and Sex in 2021, per 100,000 People, dashed lines indicate the 95% upper and lower uncertainty intervals, respectively; (C) Percentage Change in Prevalence of Renal Anemia from 1990 to 2021 and (D) Percentage Change in YLDs (per 100,000 population) during the same timeframe.

Over the past 30 years, the prevalence and YLDs rates of renal anemia in almost all age groups for both males and females have shown a declining trend, with the decline in males generally being greater than in females. However, across all age groups, the prevalence and YLDs rates for both genders are significantly higher than they were 30 years ago (figure 1C, D) (Supplementary Table S3, 4).

### (2) Geographic Trends in Renal Anemia

We found significant geographic variations in the burden of renal anemia. In 2021, the age-standardized prevalence rates (ASPR) of renal anemia were higher in Central Asia, South Asia, Southeast Asia, and Western Sub-Saharan Africa. In contrast, the ASPR of renal anemia was lower in high-income regions such as Europe and North America. Among the countries, Nepal (2,298.92/100,000; 95% UI: 1,904.13-2,765.01), Uzbekistan (1,834.86/100,000; 95% UI: 1,542.38-2,205.31), and Azerbaijan (1,759.876/100,000; 95% UI: 1,469.43-2,127.35) reported the highest ASPR. Conversely, Iceland (225.2/100,000; 95% UI: 210.36-314.62), Canada (274.56/100,000; 95% UI: 224.78-340.89), and France (290.97/100,000; 95% UI: 238.01-362.81) had the lowest ASPR (figure 2A) (Supplementary Table S5).

**Figure 2.**
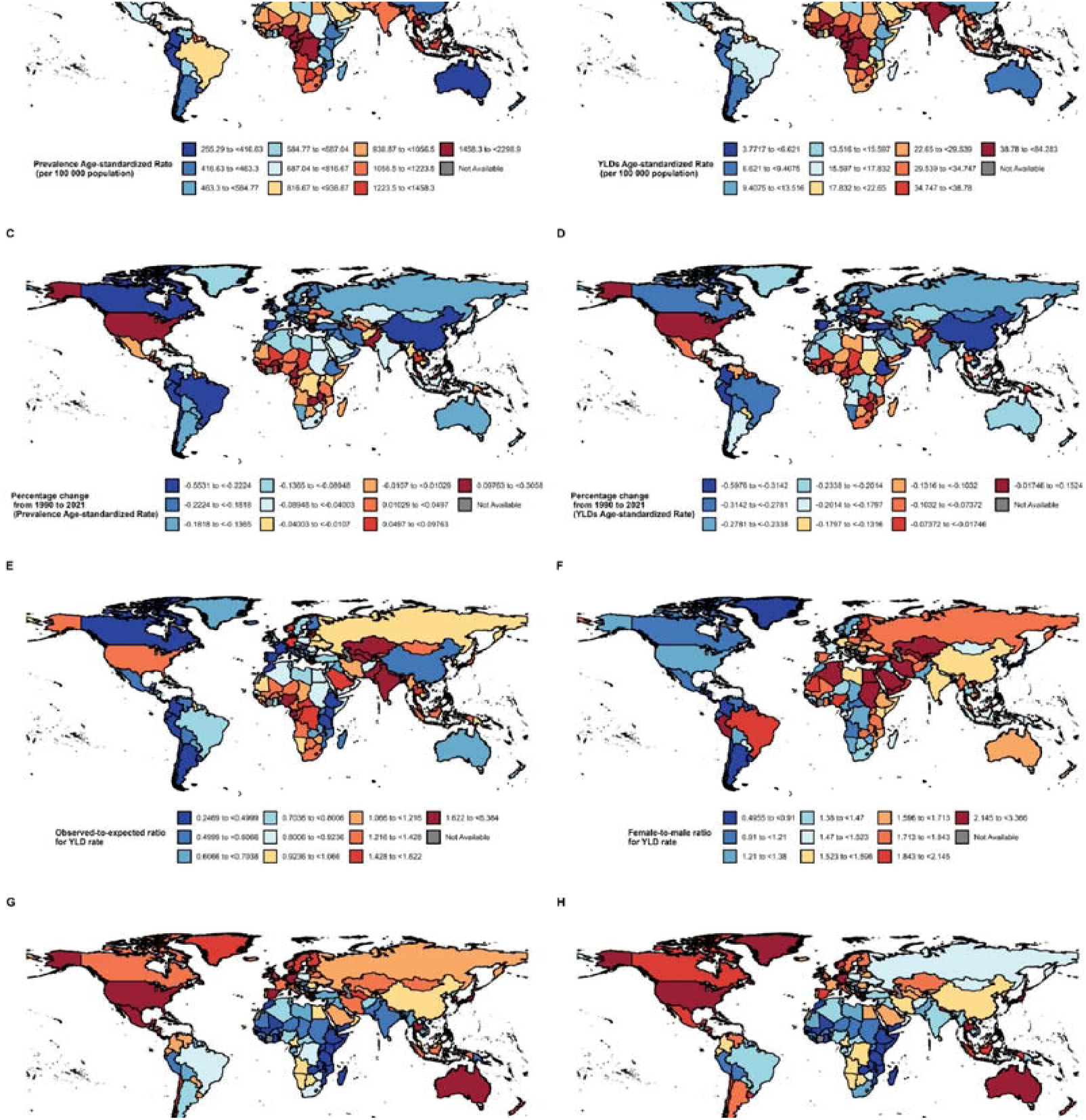
Global Burden of Renal Anemia in 2021. (A) Age-standardized Prevalence Rates (ASPR) of Renal Anemia per 100,000 Individuals in 2021; (B) Age-standardized Years Lived with Disability (YLDs) Rates of Renal Anemia per 100,000 Individuals in 2021; (C) ASPR Change from 1990 to 2021; (D) Age-Standardized YLDs Rates Change from 1990 to 2021; (E) Comparison of Observed Age-Standardized YLDs Rates versus Expected Age-Standardized YLDs Rates per 100,000 in 2021. Expected Rates Based on the Socio-demographic Index (SDI); (F) Gender Ratio of Age-Standardized YLDs Rates per 100,000 in 2021; (G) Proportion of Renal Anemia ASPR in Total Anemia ASPR; (H) Share of Renal Anemia Age-Standardized YLDs Rates in Total Anemia Age-Standardized YLDs Rates.

The geographic distribution of age-standardized YLDs rates of renal anemia in 2021 was largely consistent with that of ASPR. Nepal (84.28/100,000; 95% UI: 54.64-123.86) had the highest Age-standardized YLDs rates, followed by Nigeria (56.22/100,000; 95% UI: 36.82-80.30) and Uzbekistan (54.75/100,000; 95% UI: 34.88-82.32). The lowest age-standardized YLDs rates were observed in Canada (3.77/100,000; 95% UI: 2.21-6.12), Iceland (3.95/100,000; 95% UI: 2.33-6.24), and Monaco (4.46/100,000; 95% UI: 2.71-7.33) (figure 2B) (Supplementary Table S6).

From 1990 to 2021, there was an increasing trend in both ASPR and age-standardized YLDs rates of renal anemia in Central Asia, South Asia, Southeast Asia, and West Sub-Saharan Africa. Fiji (30.58%, 95% CI: 7.59%-57.06%), Benin (25.47%, 95% CI: 5.50%-49.97%), and Nepal (25.46%, 95% CI: 0.95%-62.67%) showed the fastest increase in ASPR. The fastest increase in age-standardized YLDs rates was observed in Fiji (15.24%, 95% CI: -0.70%-34.20%), the United States of America (14.80%, 95% CI: 1.31%-31.03%), and American Samoa (10.56%, 95% CI: -5.15%-29.14%). These trends align with the regions showing high prevalence and YLDs rates of renal anemia as illustrated in Figures 2A and 2B. Interestingly, despite the relatively low disease burden in 2021, the United States exhibited a high growth rate in ASPR (18.28%, 95% CI: 2.15%-37.66%) and age-standardized YLDs rates. In contrast, East Asia, Europe, and other parts of the Americas demonstrated a significant decline in the age-standardized prevalence and YLDs rates of renal anemia over the past 30 years (figures 2C, D) (Supplementary Table S7, 8).

Similarly, the regions with a heavy burden of renal anemia, such as Central Asia, South Asia, Southeast Asia, and West Sub-Saharan Africa, exhibited higher observed-to-expected ratios of age-standardized YLDs rates. In contrast, East Sub-Saharan Africa and South America had lower ratios (figure 2E) (Supplementary Table S9).

In 2021, across the majority of regions, the age-standardized YLDs rates of renal anemia were higher in females than in males, with the most significant gender disparity observed in Central Asia, the Middle East, North Africa, and Peru in South America (figure 2F) (Supplementary Table S10).

In high-development countries such as Japan, the United States, Singapore, Switzerland, and Australia, the age-standardized prevalence and YLDs rates of renal anemia are among the highest in the world relative to the overall age-standardized prevalence and YLDs rates of anemia. Conversely, Sub-Saharan Africa has the lowest rates (figures 2G, H) (Supplementary Table S11, 12).

### (3) Epidemiological Changes in Renal Anemia

At the regional level defined by the SDI, there is a generally negative correlation between SDI and the age-standardized prevalence rates (ASPR) and YLDs rates of renal anemia (figure S1) (Supplementary Table S13).

### (4) Rank Changes in Renal Anemia

Compared to 1990, the ASPR ranking of renal anemia (anemia caused by chronic kidney disease) in 2021 increased by one position in Southeast Asia, South Asia, and Oceania, and by three positions in Sub-Saharan Africa (figure S2A, B) (Supplementary Table S14). Meanwhile, the age-standardized YLDs rates ranking of renal anemia in 2021 increased by one position globally and by three positions in Sub-Saharan Africa (figure S3C, D) (Supplementary Table S15).

### (5) Cross-country inequality analysis of renal anemia age-standardized prevalence and YLDs

Significant absolute and relative SDI-related inequalities were observed in renal anemia, with countries having lower SDI bearing higher age-standardized YLDs rates and ASPR. As indicated by the inequality slope index, the gap in ASPR between the highest and lowest SDI countries increased from -264.65 in 1990 to -451.16 in 2021 (figure 3A) (Supplementary Table S16), and the gap in age-standardized YLDs rates increased from -23.62 in 1990 to -24.13 in 2021 (figure 3C) (Supplementary Table S17). In the inequality concentration curves, both in 1990 and 2021, the curves are above the equality line, indicating that the main contributors to age-standardized YLDs rates and ASPR of renal anemia are low SDI regions. Although the 2021 curve is closer to the equality line compared to 1990, the improvement is very limited (figure 3B, D) (Supplementary Table S16, 17).

**Figure 3.**
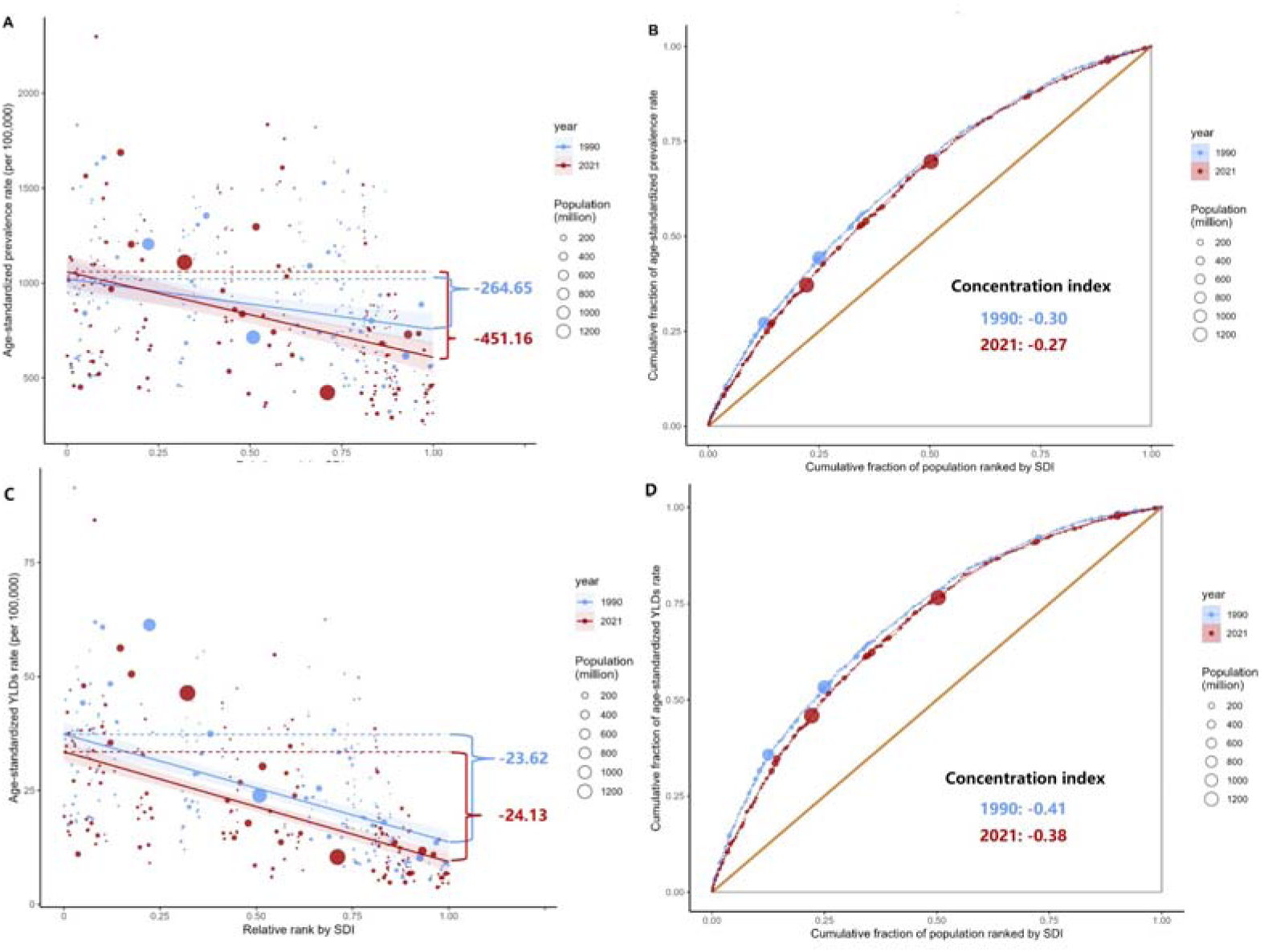
Health Inequity Analysis for Renal Anemia. (A) Health Inequity Regression Curve and (B) Concentration Curve for ASPR of Renal Anemia. (C) Health Inequity Regression Curve and (B) Concentration Curve for Age-Standardized YLDs Rates of Renal Anemia.

### (6) Frontier Analysis on the Basis of Age-Standardized YLDs

In low to middle SDI regions, there is no significant difference in the effective fraction (EF) of ASPR corresponding to specific SDI levels as SDI increases. However, in middle to high SDI regions, the EF of ASPR decreases as SDI increases (figure S4A) (Supplementary Table S18). This trend is more evident in the frontier analysis of age-standardized YLDs rates (figure 4A) (Supplementary Table S19). The five countries with the highest EF of ASPR are Nepal, Uzbekistan, Azerbaijan, Thailand, and the Democratic Republic of the Congo (figure S4B) (Supplementary Table S18); whereas the five countries with the largest difference between age-standardized YLDs rates and expected values are Nepal, Nigeria, Pakistan, India, and the Democratic Republic of the Congo (figure 4B) (Supplementary Table S19).

### (7) Age-Period-Cohort Model Analysis

In regions classified based on SDI, the age effect pattern is the same, with the risk of disease increasing with age. Among these, the prevalence is highest in elderly individuals in high-middle SDI regions, while it is lowest in elderly individuals in low SDI regions (figure S5A) (Supplementary Table S20). In terms of period and birth cohort effects, the prevalence in all SDI regions shows a general downward trend, with the fastest decline in high-middle SDI regions, while the prevalence declines more slowly in low-middle SDI and low SDI regions (figure S5B, C) (Supplementary Table S20).

**Figure 4.**
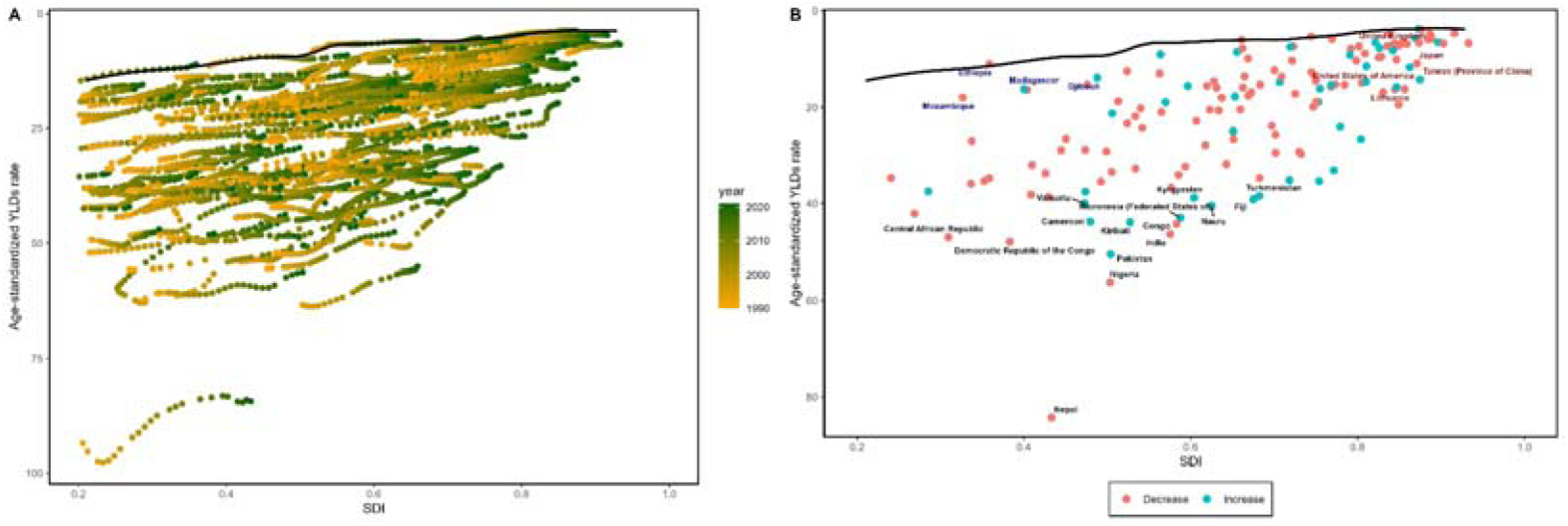
Frontier Analysis of Age-Standardized YLDs Rates of Renal Anemia Based on SDI. (A) Frontier Analysis for SDI and Age-Standardized YLDs Rates for the Period 1990-2021; (B) Frontier Analysis for Different Countries Based on SDI and Age-Standardized YLDs Rates in 2021. The frontier line is delineated in black, indicating the potentially achievable ASPR or age-standardized YLDs on the basis of SDI; dots represent the actual ASPR or age-standardized YLDs in every country and territory. The top fifteen countries with the highest effective difference are labeled in black. The top countries with the lowest effective difference in low SDI are labeled in blue, whereas the highest effective difference in very high SDI are labeled in red.

For YLDs analysis, the age effect also shows an increasing trend in YLDs rates with age. Unlike prevalence, elderly individuals in low-middle and low SDI regions have higher YLDs rates (figure 5A) (Supplementary Table S21). In terms of period and birth cohort effects, YLDs rates overall show a downward trend, with the fastest decline also observed in high-middle SDI regions (figure 5B, C) (Supplementary Table S21).

**Figure 5.**
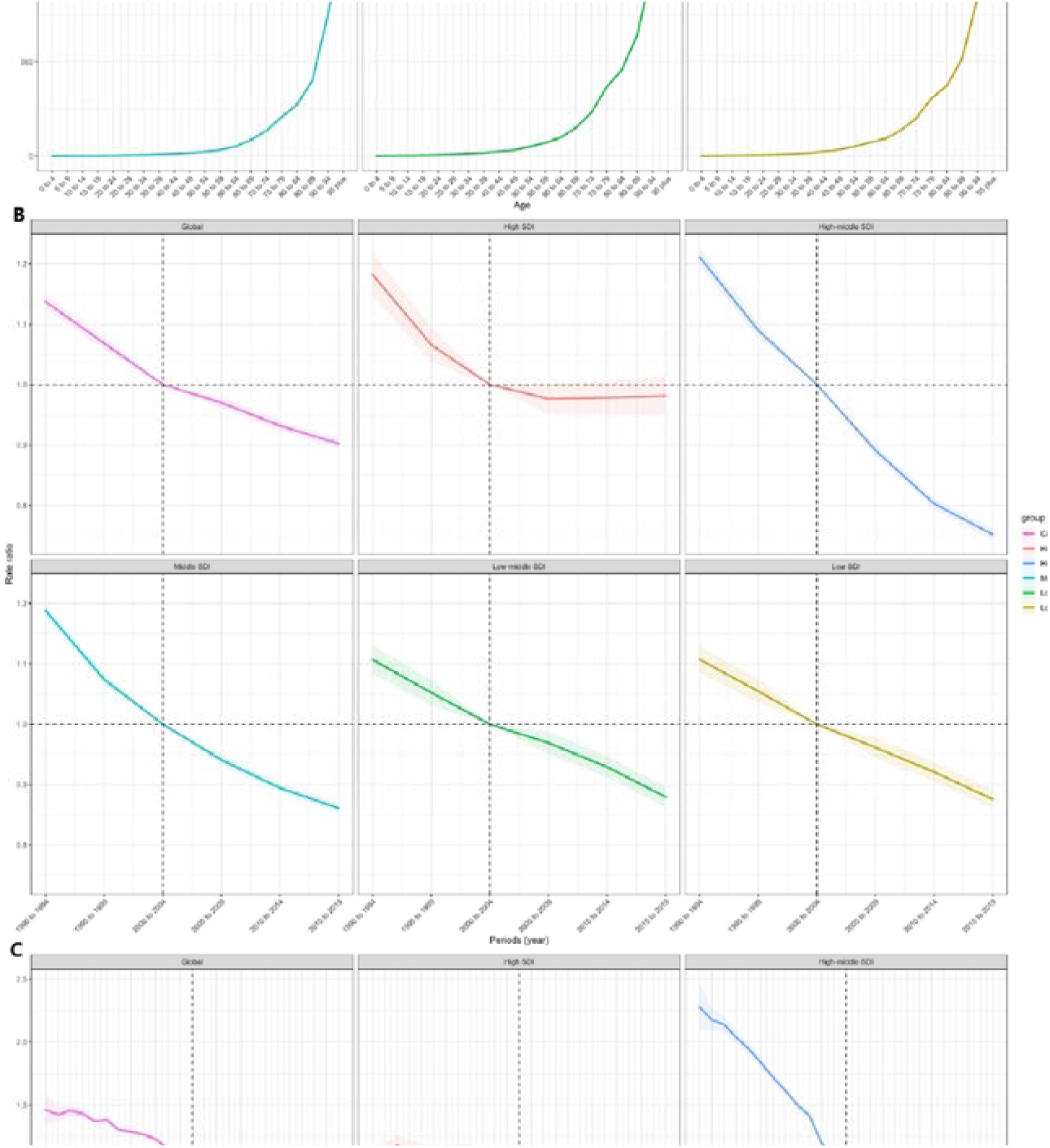
Age-Period-Cohort Analysis for the YLDs of Renal Anemia Based on SDI. (A) Age Effect (B) Period Effect (C) Cohort Effect.

## Discussion

In 2021, over a quarter of the global population was afflicted with anemia, contributing to 5.7% of the world’s Years Lived with Disability (YLDs). Although the prevalence of anemia has been gradually declining in recent years^[9]^, the current rates of anemia and related YLDs remain substantial and cannot be overlooked. Amidst a general decrease in the burden of anemia, the proportion attributable to chronic kidney disease has been on the rise, indicating that improvements in anemia burdens associated with CKD lag behind overall progress. Thus, for the first time, this study has analyzed the burden of renal anemia using 2021 data at global, regional, and national levels to fill this research gap.

Our findings reveal that individuals over the age of 65 bear the greatest burden of renal anemia. This age group is particularly susceptible to CKD-induced renal anemia, as CKD patients cannot combat the disease effectively due to low EPO production^[15]^. A 2023 observational study in the United States corroborates these findings, involving over five million patients with an average age of fifty-four, concluding that the prevalence and severity of anemia significantly increases as the eGFR decreases^[15]^. Such pronounced anemia severity is closely linked to elevated risks of cardiovascular diseases, coronary artery disease, stroke, heart failure, and increased mortality^[15]^. Although the mechanisms remain unspecified, several studies demonstrate that iron deficiency is common in renal anemia^[4,15]^. Consequently, iron supplements are prioritized in renal anemia guidelines in many countries^[10]^, while erythropoiesis-stimulating agents (ESAs) are not usually the first choice due to potential risks of stroke, hypertension, and tumors^[10]^. In 2018, the hypoxia-inducible factor prolyl hydroxylase inhibitor (HIF-PHI) Roxadustat was approved for treatment of renal anemia in China. Japan, the European Union, India, and the United States subsequently approved Roxadustat or other HIF-PHIs for renal anemia treatment. Clinical research also verifies the efficacy of Roxadustat in treating anemia in both dialysis^[16]^ and non-dialysis CKD patients^[17]^. This is an encouraging shift, yet globally, renal anemia still does not receive high levels of attention, and nations should fortify support for the prevention and treatment of the condition.

In 2021, Central Asia, South Asia, Southeast Asia, and Western Sub-Saharan Africa bore a heavy burden of renal anemia. Similarly, these regions experienced the fastest growth in the burden of renal anemia from 1990 to 2021. Due to the lack of early diagnosis and standardized treatment, these regions face a heavy burden of hypertension^[18,19]^, which in turn exacerbates the incidence of CKD complications^[13,20]^. Additionally, CKD patients in these regions are unable to receive adequate treatment^[21,22]^, leading to a quicker progression to end-stage kidney disease (ESKD). In contrast, high-income regions such as Europe and North America have more comprehensive healthcare systems, resulting in a lighter burden of renal anemia and slower growth rates. The only exception is the United States, where a relatively low disease burden is accompanied by a very high burden growth rate. This is mainly attributed to the exceptionally high obesity growth rate in the U.S. compared to other developed countries^[23,24]^. The higher socioeconomic development level in the U.S. allows people to consume more high-sugar, high-fat diets, and the industrialization process has led to reduced physical activity, all contributing to increased obesity rates. High obesity rates further increase the incidence of chronic diseases such as diabetes^[25]^ and hyperlipidemia^[26]^, most of which eventually complicate with chronic kidney disease^[27,28]^.

At the regional level, there is a negative correlation between the level of SDI and the age-standardized prevalence rates (ASPR) and age-standardized YLDs rates of renal anemia. As previously mentioned, residents of high-SDI regions can access higher quality medical resources^[29,30]^, resulting in lower incidences of hypertension, diabetes, and hyperlipidemia, which are precursor diseases to CKD, ultimately leading to a lighter burden of renal anemia. Despite the exception of the United States, this does not significantly affect the global trend. According to the standard health equity analysis methods recommended by the WHO, we conducted a cross-national inequality analysis of renal anemia^[12]^, which showed that countries with lower SDI bear higher burden of renal anemia. Moreover, this inequality has been increasing over time. Frontier analysis shows that there is greater room for improvement in the ASPR and age-standardized YLDs rates of renal anemia in low and middle SDI regions compared to their expected values. The age-period-cohort model analysis indicates that the burden of renal anemia has been reducing slowly over the past 30 years in low-middle and middle SDI regions. These findings suggest that attention to renal anemia and the allocation of related medical resources in underdeveloped regions remain insufficient.

Compared to previous GBD publications on anemia, the chief advantage of our current study is that we provide deeper insight into the status and trends of renal anemia, a subset of anemia across populations, time, and space—offering valuable insights for more precise epidemiological and health policy implementations. Specifically, the latest GBD 2021 results on anemia show heavy burdens among children, women of reproductive age, and the elderly, particularly in underdeveloped areas like Sub-Saharan Africa, Central Asia, and South Asia^[9]^. Existing health policies globally are largely aware of these results and have made targeted efforts to address anemia in women of reproductive age and children in underdeveloped regions^[31,32]^. Accordingly, over the past thirty years, the burden of anemia has indeed been diminishing not only in these areas but globally^[33]^, yet can we not do better? In these regions, there are still many elderly people suffering from renal anemia. In the developed country of the United States, the burden of renal anemia is significantly increasing. Providing targeted prevention, diagnosis, and treatment for renal anemia to the elderly populations in these regions would yield greater benefits from the limited medical resources. Despite these advantages, our study has limitations. Firstly, due to the lower healthcare standards in some underdeveloped countries, misdiagnosis and underdiagnosis may lead to underestimation of disease prevalence and YLDs. Secondly, current data do not include data on renal anemia caused by CKD at various stages, and we hope to supplement these data in the future. Thirdly, substantial reliance on statistical modeling methods used by GBD collaborators, particularly at the country level, means data sourced from GBD heavily depends on modeled data due to lack of raw data. Fourthly, the absence of sub-national data precludes exploration of sub-national trends. Lastly, the latency of GBD data is also a non-negligible issue.

Anemia is a principal public health concern worldwide, with substantial inter-country variations, and different nations have different primary types of anemia; uniform healthcare policies might fall short of efficiency. While the age-standardized prevalence and YLDs of anemia have declined from 1990 to 2021, the escalating global aging population means renal anemia associated with older chronic diseases will undoubtedly constitute a larger share of the anemia burden in the future. Tackling the burden of renal anemia calls for comprehensive intervention measures, with priority consideration for the highest risk groups—mainly the elderly in middle and low development regions, as well as the obese population in the United States. Strengthening early diagnosis and treatment of CKD on one hand, and advocating for low sugar and low-fat diets to relieve the burden of diabetes and hyperlipidemia, chronic diseases inducing CKD. On the other hand, the development of more effective and low-side-effect renal anemia treatments should not be overlooked.

## Contributors

HZ conceived the study and designed the protocol with FC. QG, LF, JJ, TM and TH analyzed the GBD data. FC, JJ and YM were responsible for the article writing. HS, HZ, and XL were responsible for the article checking and reviewing. HS, XL, and QG accessed and verified the data. HZ had access to all the data in the study and had final responsibility for the decision to submit for publication.

## Data sharing

The data used in this study are freely available for download from the GBD 2021 website (https://vizhub.healthdata.org/gbd-results/). All data used in this study will also be made available on request to the corresponding author.

## Data Availability

All data produced in the present study are available upon reasonable request to the authors
All data produced in the present work are contained in the manuscript
All data produced are available online at: https://vizhub.healthdata.org/gbd-results/

## Acknowledgments

We acknowledge the Institute for Health Metrics and Evaluation, the GBD Diseases and Injuries Collaborators, and all staff who shared their data needed for this study.

This work was supported by the National Natural Science Foundation of China [No. 82074166], Huadong Medicine Joint Funds of the Zhejiang Provincial Natural Science Foundation of China [No. LHDMZ24H050001], Fujian Provincial Natural Science Foundation [No. 2018J01121], Fujian Provincial Health Technology Project [No. 2020GGA026], and Medical Discipline Construction Project of Pudong Health Committee of Shanghai [No. PWYts2021-18].

## References

[1] Macdougall IC. Anaemia in CKD-treatment standard. Nephrology Dialysis Transplantation. 2023:gfad250.

[2] Souma T, Yamazaki S, Moriguchi T, Suzuki N, Hirano I, Pan X, et al. Plasticity of renal erythropoietin-producing cells governs fibrosis. Journal of the American Society of Nephrology. 2013;24(10):1599–616.

[3] Locatelli F, Del Vecchio L. Quality of life: a crucial aspect for the patients, a neglected goal in the treatment of anemia in patients with CKD. Kidney International. 2023;103(6):1025–7.

[4] Farrington DK, Sang Y, Grams ME, Ballew SH, Dunning S, Stempniewicz N, et al. Anemia prevalence, type, and associated risks in a cohort of 5.0 million insured patients in the United States by level of kidney function. American Journal of Kidney Diseases. 2023;81(2):201–9. e1.

[5] Glassock RJ, Warnock DG, Delanaye P. The global burden of chronic kidney disease: estimates, variability and pitfalls. Nature Reviews Nephrology. 2017;13(2):104–14.

[6] Inker LA, Grams ME, Levey AS, Coresh J, Cirillo M, Collins JF, et al. Relationship of estimated GFR and albuminuria to concurrent laboratory abnormalities: an individual participant data meta-analysis in a global consortium. American Journal of Kidney Diseases. 2019;73(2):206–17.

[7] Li Y, Shi H, Wang W-M, Peng A, Jiang G-R, Zhang J-Y, et al. Prevalence, awareness, and treatment of anemia in Chinese patients with nondialysis chronic kidney disease: first multicenter, cross-sectional study. Medicine. 2016;95(24):e3872.

[8] UNICEF. The extension of the 2025 maternal, infant and young child nutrition targets to 2030: WHO. 2021.

[9] Collaborators GA. Prevalence, years lived with disability, and trends in anaemia burden by severity and cause, 1990–2021: findings from the Global Burden of Disease Study 2021. The Lancet Haematology. 2023;10(9):e713-e34.

[10] Mikhail A, Brown C, Williams JA, Mathrani V, Shrivastava R, Evans J, et al. Renal association clinical practice guideline on Anaemia of Chronic Kidney Disease. BMC nephrology. 2017;18:1–29.

[11] Kliger AS, Foley RN, Goldfarb DS, Goldstein SL, Johansen K, Singh A, et al. KDOQI US commentary on the 2012 KDIGO clinical practice guideline for anemia in CKD. American Journal of Kidney Diseases. 2013;62(5):849–59.

[12] Organization WH. Handbook on health inequality monitoring: with a special focus on low-and middle-income countries: World Health Organization; 2013.

[13] Xie Y, Bowe B, Mokdad AH, Xian H, Yan Y, Li T, et al. Analysis of the Global Burden of Disease study highlights the global, regional, and national trends of chronic kidney disease epidemiology from 1990 to 2016. Kidney International. 2018;94(3):567–81.

[14] Cao F, Li D-P, Wu G-C, He Y-S, Liu Y-C, Hou J-J, et al. Global, regional and national temporal trends in prevalence for musculoskeletal disorders in women of childbearing age, 1990–2019: an age-period-cohort analysis based on the Global Burden of Disease Study 2019. Annals of the Rheumatic Diseases. 2024;83(1):121-32.

[15] Stauder R, Valent P, Theurl I. Anemia at older age: etiologies, clinical implications, and management. *Blood*, The Journal of the American Society of Hematology. 2018;131(5):505–14.

[16] Chen N, Hao C, Liu B-C, Lin H, Wang C, Xing C, et al. Roxadustat treatment for anemia in patients undergoing long-term dialysis. New England Journal of Medicine. 2019;381(11):1011–22.

[17] Chen N, Hao C, Peng X, Lin H, Yin A, Hao L, et al. Roxadustat for anemia in patients with kidney disease not receiving dialysis. New England Journal of Medicine. 2019;381(11):1001–10.

[18] Mills KT, Stefanescu A, He J. The global epidemiology of hypertension. Nature Reviews Nephrology. 2020;16(4):223–37.

[19] Zhou B, Perel P, Mensah GA, Ezzati M. Global epidemiology, health burden and effective interventions for elevated blood pressure and hypertension. Nature Reviews Cardiology. 2021;18(11):785–802.

[20] Bikbov B, Purcell CA, Levey AS, Smith M, Abdoli A, Abebe M, et al. Global, regional, and national burden of chronic kidney disease, 1990–2017: a systematic analysis for the Global Burden of Disease Study 2017. The Lancet. 2020;395(10225):709-33.

[21] Ashuntantang G, Osafo C, Olowu WA, Arogundade F, Niang A, Porter J, et al. Outcomes in adults and children with end-stage kidney disease requiring dialysis in sub-Saharan Africa: a systematic review. The Lancet Global Health. 2017;5(4):e408–e17.

[22] Tang SC, Yu X, Chen HC, Kashihara N, Park HC, Liew A, et al. Dialysis care and dialysis funding in Asia. American Journal of Kidney Diseases. 2020;75(5):772–81.

[23] Blüher M. Obesity: global epidemiology and pathogenesis. Nature Reviews Endocrinology. 2019;15(5):288–98.

[24] Topór-Mądry R. Health Effects of Overweight and Obesity in 195 Countries over 25 Years. New England Journal of Medicine. 2017;377(1).

[25] Ong KL, Stafford LK, McLaughlin SA, Boyko EJ, Vollset SE, Smith AE, et al. Global, regional, and national burden of diabetes from 1990 to 2021, with projections of prevalence to 2050: a systematic analysis for the Global Burden of Disease Study 2021. The Lancet. 2023;402(10397):203–34.

[26] Repositioning of the global epicentre of non-optimal cholesterol. Nature. 2020;582(7810):73-7.

[27] Webster AC, Nagler EV, Morton RL, Masson P. Chronic kidney disease. The Lancet. 2017;389(10075):1238–52.

[28] Romagnani P, Remuzzi G, Glassock R, Levin A, Jager KJ, Tonelli M, et al. Chronic kidney disease. Nature reviews Disease primers. 2017;3(1):1–24.

[29] Vanholder R, Annemans L, Braks M, Brown EA, Pais P, Purnell TS, et al. Inequities in kidney health and kidney care. Nature Reviews Nephrology. 2023;19(11):694–708.

[30] Love-Koh J, Griffin S, Kataika E, Revill P, Sibandze S, Walker S. Methods to promote equity in health resource allocation in low-and middle-income countries: an overview. Globalization and health. 2020;16:1–12.

[31] Kapil U, Kapil R, Gupta A. National Iron Plus Initiative: current status & future strategy. Indian Journal of Medical Research. 2019;150(3):239–47.

[32] Tiruneh FN, Asres DT, Tenagashaw MW, Assaye H. Decision-making autonomy of women and other factors of anemia among married women in Ethiopia: a multilevel analysis of a countrywide survey. BMC public health. 2021;21(1):1497.

[33] Safiri S, Kolahi A-A, Noori M, Nejadghaderi SA, Karamzad N, Bragazzi NL, et al. Burden of anemia and its underlying causes in 204 countries and territories, 1990–2019: results from the Global Burden of Disease Study 2019. Journal of hematology & oncology. 2021;14:1-16.

