## Supplementary images for "Burden of renal anemia in 204 countries and territories, 1990-2021 : results from the Global Burden of Disease Study 2021" (from another version of this preprint: version 3, 2024-07-02)

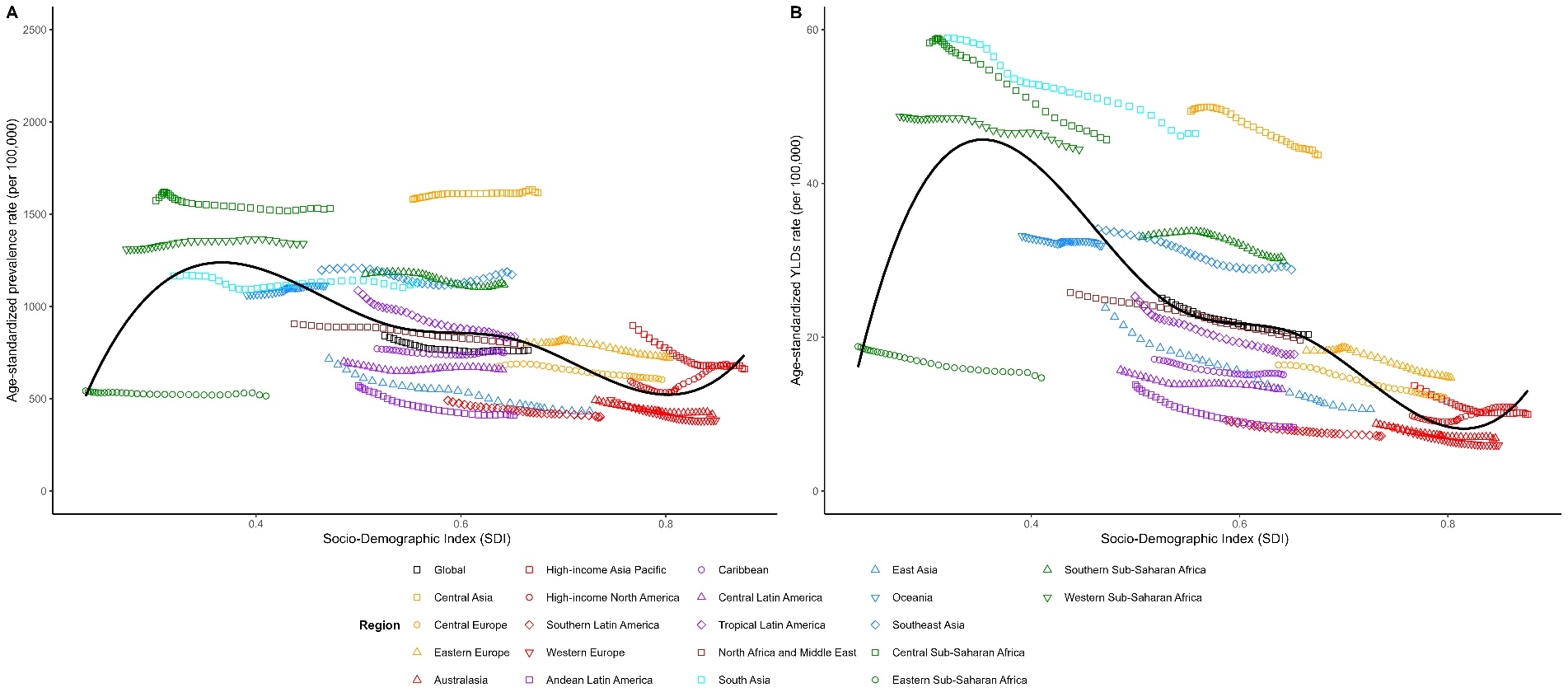

**Figure S1. Age-Standardized Prevalence and YLDs rates of Renal Anemia in 21 Global Disease Burden Regions Stratified by SDI, 1990-2021** (A) ASPR of Renal Anemia from 1990 to 2021, in 21 Global Burden of Disease (GBD) Regions Categorized by the SDI; (B) Age-Standardized YLDs Rates for Renal Anemia from 1990 to 2021, in the Same GBD Regions Arranged by the SDI).

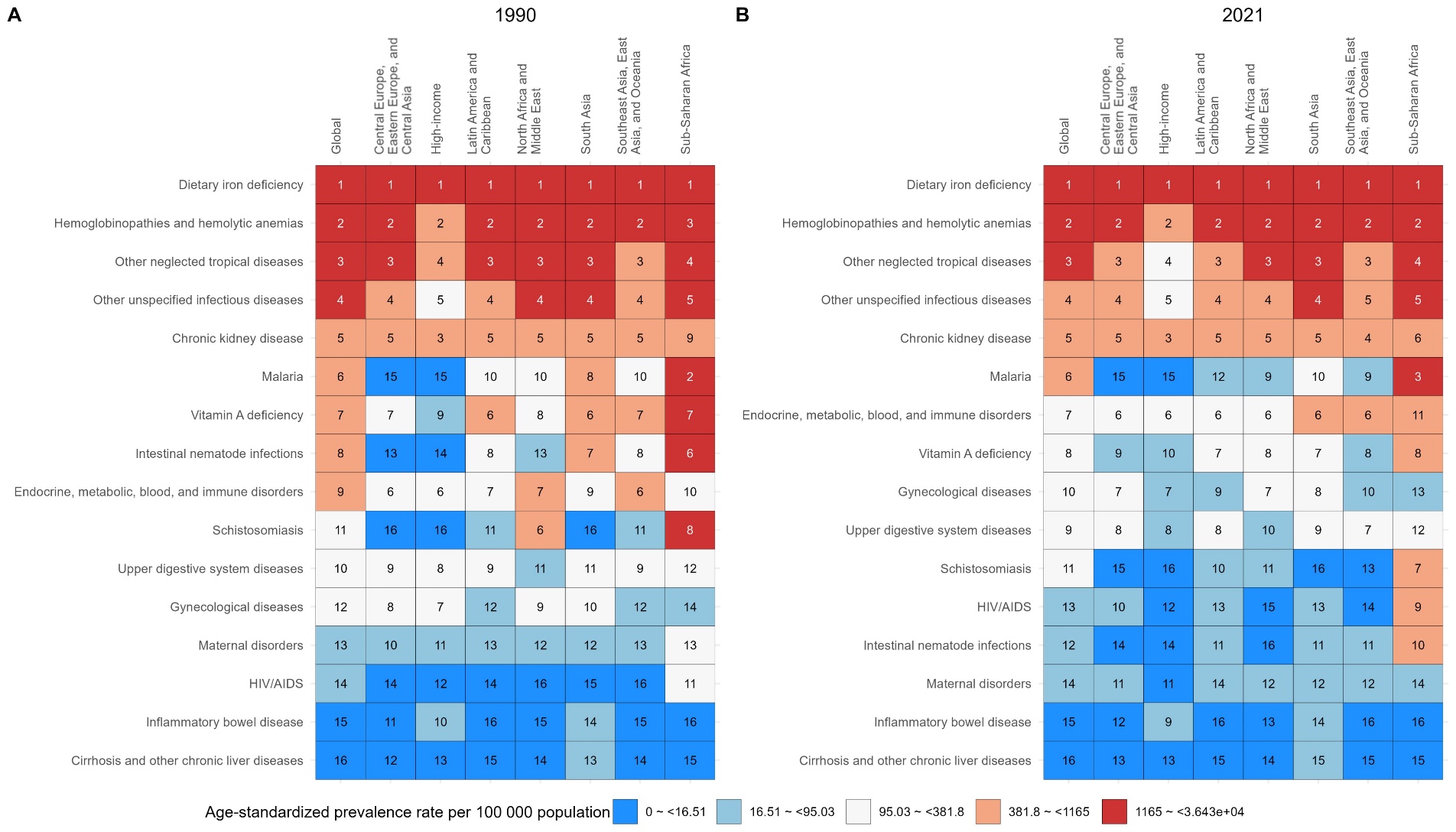

**Figure S2. Ranking of Reasons for ASPR of Renal Anemia** (A) Global and Super-Regional Ranking of Renal Anemia Causes by ASPR Across All Age Groups in 1990 and (B) in 2021.

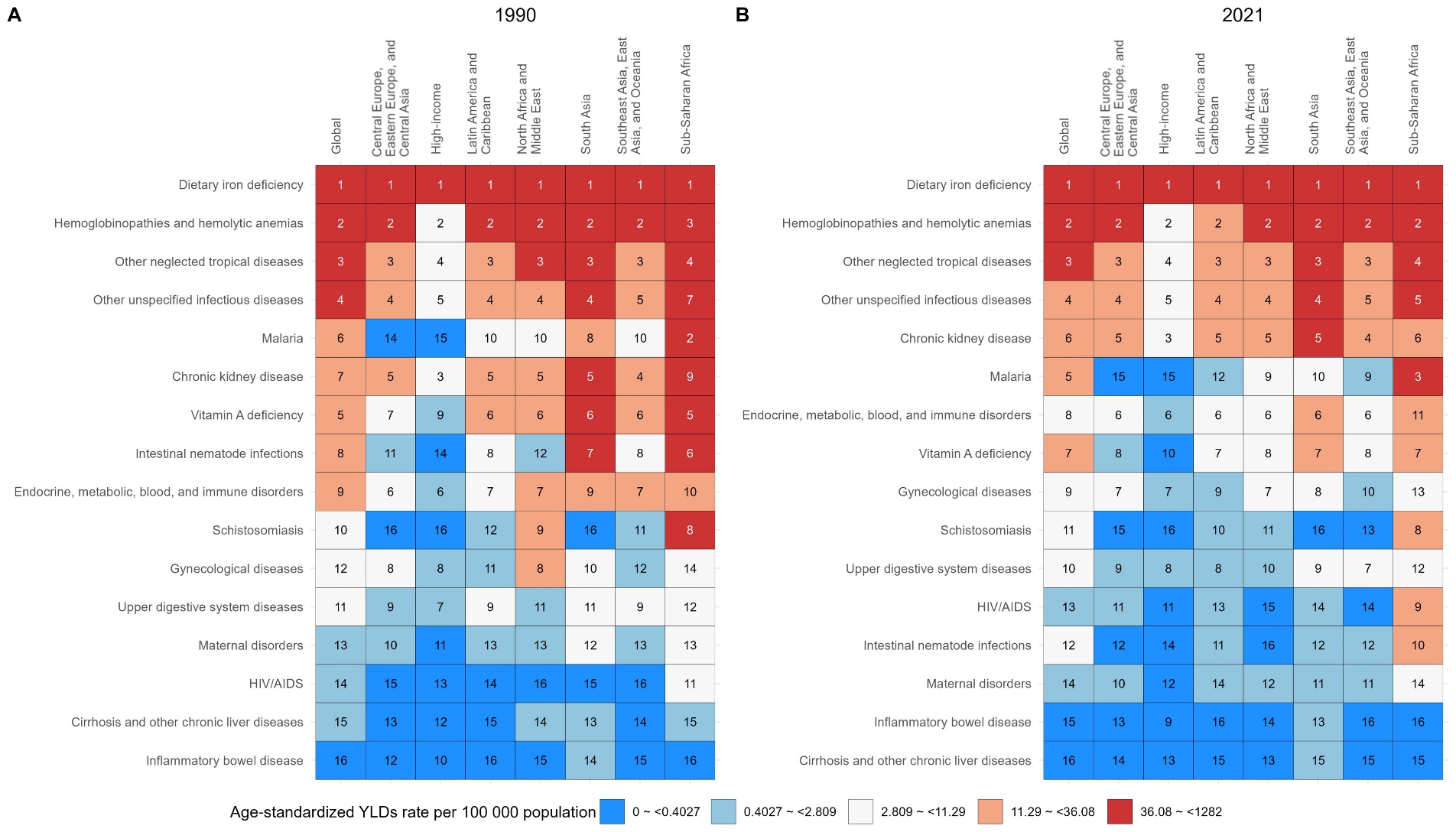

**Figure S3. Ranking of Reasons for Age-Standardized YLDs Rates of Renal Anemia** (A) Global and Super-Regional Ranking of Renal Anemia Causes by Age-Standardized YLDs Rates Across All Age Groups in 1990 and (B) in 2021.

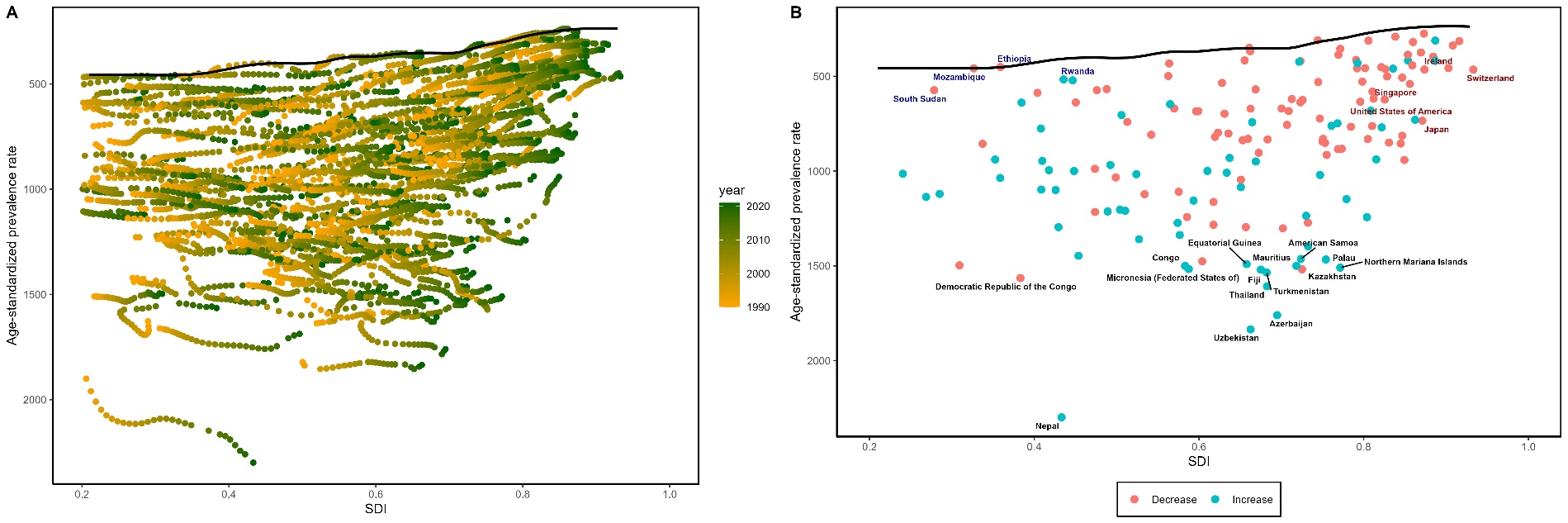

**Figure S4. Frontier Analysis of ASPR of Renal Anemia Based on SDI** (A) Frontier Analysis for SDI and ASPR for the Period 1990-2021; (B) Frontier Analysis for Different Countries Based on SDI and ASPR in 2021.

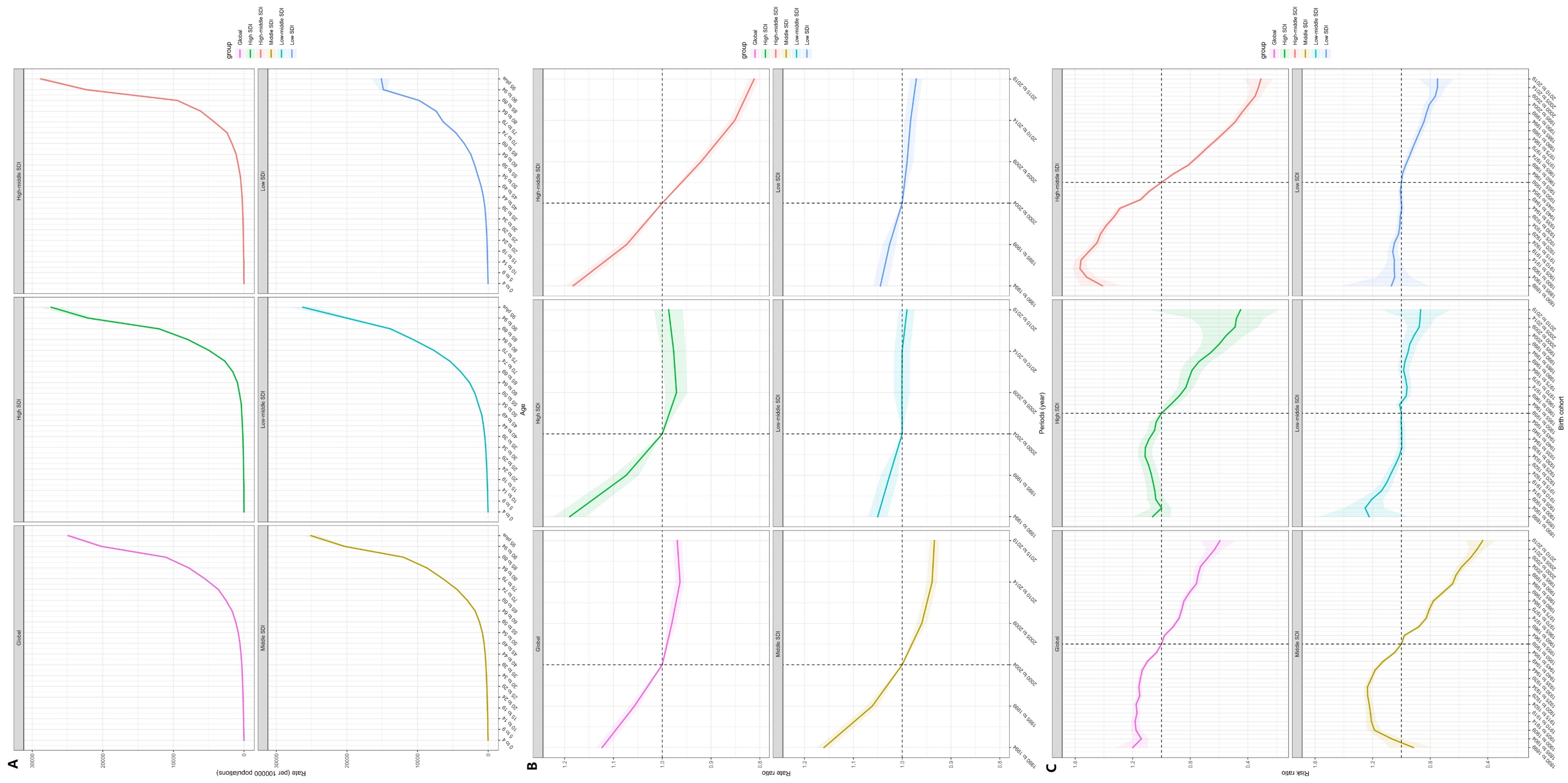

**Figure S5. Age-Period-Cohort Analysis for the Prevalence of Renal Anemia Based on SDI** (A) Age Effect (B) Period Effect (C) Cohort Effect.
